# Cerebral Blood Flow with multi-delay Arterial Spin Labeling MRI Reveals Interictal Hypo- and Hyper-perfusion in epilepsy: A Proof of Concept

**DOI:** 10.64898/2026.01.21.26344148

**Authors:** Myriam Abdennadher, Lena Václavů, Joseph Sisto, Shreeya Patel, Leonie Petitclerc, Meher R Juttukonda, Muhammad M Qureshi, Abrar Al-Faraj, Maria Stefanidou, Janine Barrett, Indrani Alagar, Sara Jacobellis, Chad Farris, Osamu Sakai, Lee E. Goldstein, Ali Guermazi, Sara K. Inati, William H. Theodore, Bruce Rosen, Matthias J.P. van Osch, Ning Hua

## Abstract

**Background and Purpose:** Interictal cerebral blood flow (CBF) may be useful for seizure focus localization. However, its accuracy is debatable. Studies suggested interictal hypoperfusion at the seizure focus, yet others suggested hyperperfusion. This study aims to investigate the patterns of interictal perfusion in epilepsy subjects compared to healthy controls using multiple labeling-delays arterial spin labeling MRI, and to explore the accuracy of perfusion estimation using single post-labeling delay arterial-spin labeling MRI by comparing it to multiple post-labeling delays arterial-spin labeling MRI.

**Materials and Methods:** We analyzed CBF in 40 participants, 15 healthy (35.9 ± 9.5 years, 47% women) and 25 epilepsy (41.1 ± 10.5 years; 52% women). We acquired a multiple post-labeling delays arterial spin labeling MRI using Hadamard encoding, and structural MRI. Perfusion quantification was performed using in-house software and analyzed using surface-based method. Z-scores of CBF and its absolute value (|Z-score|) were calculated to evaluate abnormal perfusion.

**Results:** Brain perfusion showed interictal hypo- and/or hyper-perfusion in epilepsy compared to healthy participants involving focal to whole brain alterations. Epilepsy subjects had higher |Z-score| in most cortical regions compared to the healthy group. CBF generated from single post-labeling delay arterial-spin labeling MRI correlated with ones from multiple post-labeling delays in most cortical regions, yet the level of correlation was affected by regional arterial transit time and the labeling scheme. In regions with shorter arterial transit time, correlation peaked at shorter post-labeling delay (1300ms); whereas in regions with longer arterial transit time, correlation peaked at longer post-labeling delay (2250ms).

**Conclusions:** Our findings suggest that the interictal CBF may fluctuate between hypo- and hyper-perfusion, and the involvement of brain regions may extend well beyond the seizure focus. Additionally, while single-post-labeling delay is efficient and clinically feasible, the accuracy of its assessment of perfusion depends on brain regions and the labeling scheme.

**SUMMARY STATEMENT:** We observed interictal hypo-/hyper-perfusion, possibly extending beyond the seizure focus. Caution is warranted when interpreting single-post-labeling-delay perfusion MRI in epilepsy, as its reliability varies by brain region and labeling scheme.

**Key Results:**

- Epilepsy subjects showed larger fluctuation of CBF, yet the fluctuation may happen in either direction: hypo- or hyper-perfusion.
- The extent of fluctuation can be focal, regional, hemispheric, or global.
- CBF derived from single-PLD MRI largely correlates with the ones derived from more reliable multi-PLD method. However, the level of correlation varies spatially (i.e., brain regions) and temporally (i.e., different labeling and post-labeling schemes), and may be explained by regional variability of arterial transit times.

## INTRODUCTION

Epilepsy is a common neurological disorders and affects 6.38 per 1000 people globally with variability across ages, genders, races, and countries (1). Focal epilepsy is the predominant type in adults and children (2, 3). Around 20-40% of epilepsy patients exhibit poor prognosis with persistent seizures despite multiple medication trials (1), many of whom may benefit from surgery (4). However, surgery success relies on adequate seizure focus localization.

Cerebral blood flow (CBF) has been explored to aid in the localization of the seizure focus or even predict surgical outcomes (5-11). The main utilization of CBF measures is to identify asymmetric perfusion patterns between brain hemispheres (8-12). Relative interictal hypo-perfusion was found in the area ipsilateral to the seizure focus, yet with questionable accuracy (6, 9, 12, 13). This is partially because the asymmetric measurement is a relative index and cannot distinguish if the subject is experiencing a global hypo- or hyper-perfusion, and seizure focus laterality in bilateral cases (depending on the test timing and last seizure).

To advance our understanding of the relationship between seizure focus and interictal CBF, we investigated patients with focal epilepsy, compared to healthy controls, using a surface-based vertex-wise analysis. In this study, we used pseudo-continuous arterial spin labeling (pCASL) MRI to measure CBF. Compared to other techniques, pCASL MRI is safe, non-invasive, and does not need radioactive tracers or exogenous contrast agents (14). While pCASL MRI using multiple post labeling delays (PLDs) generates more reliable CBF measurement (14), single PLD pCASL MRI (typically shorter) has been in clinical use due to its widespread availability. Therefore, we also examined the deviation patterns of single PLD from multiple-PLDs in measuring CBF in our study cohort.

## MATERIALS AND METHODS

### Subject Recruitment

The study was approved by the Institutional Review Board. Forty-one participants were enrolled between September 2022 and January 2024. One subject was excluded from this analysis due to surface segmentation error. Forty were included in this study: 15 healthy and 25 epilepsy individuals. The inclusion criteria were: focal epilepsy and 18 years or older. The exclusion criteria were: pregnancy or any MRI contraindication; brain trauma; stroke/vascular malformation; brain tumor/surgery; vagus nerve stimulator; heart failure; bilateral tonic-clonic seizure within 24 hours from visit; and limited MRI data quality (artifact, segmentation error, etc.). Fifteen sex- and age-matched healthy subjects without known neurological or psychiatric conditions and no active recreational substance use were recruited. Structural MRI data were reviewed by two neuroradiologists (OS, CF, 21 and 4 years of experience respectively).

### MRI Data Acquisition

Brain MRI data were acquired using a 3T Philips MR 7700 imaging system with a 32-channel head coil. We obtained 3D T1-weighted structural images. Key parameters were: flip angle=8°, repetition time=6.7ms, echo time=3.0ms, field of view=240×240×170mm^3^, acquired resolution=1×1×1mm^3^. We used a pCASL sequence with a Hybrid Hadamard-4 labeling scheme(15) and a 3D gradient and spin-echo readout. The labeling durations (LD) were 650ms, 650ms, 950ms, 650ms, 950ms, 950ms, 2000ms, 2000ms, 2000ms, and the corresponding post-labelling delays for each LD were 200ms, 650ms, 850ms, 900ms, 1300ms, 1550ms, 1800ms, 2250ms 2500ms. Other key parameters: repetition time=6000ms, echo time=12ms, field of view=240×240×108 mm^3^, voxel resolution = 3.75×3.75×6mm^3^. A separate equilibrium magnetization (M0) image for voxel-wise calibration was acquired with the same geometry settings as the ASL scan. Key parameters were repetition time=2000ms, echo time=12ms.

### MRI Data Analysis

CBF was calculated via both single PLD and multiple PLD methods using in-house software programmed in MATLAB (MathWorks, MA). For each single PLD, the CBF was quantified according to recommendations from the ISMRM Perfusion Study Group (14).

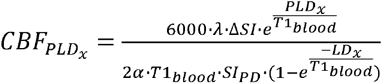

In which, λ is the blood-brain partition coefficient of 0.9 ml/g (16); ⍰ is the labeling efficiency 0.85 (17); T1_blood_ is the T1 of arterial blood assumed to equal 1650ms; ΔSI is the signal difference equal to the subtracted ASL data; SI_PD_ is TR-adjusted signal intensity from the separately acquired proton density images. The multiple PLD derived CBF (CBF_multi_) is calculated using the method summarized by Woods et. al.(18) and described in the equation below.

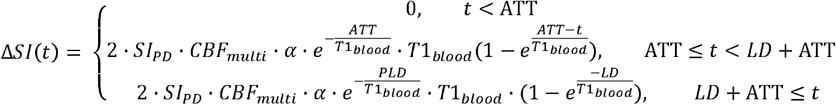

Therefore, for multiple-PLD, the ATT maps were also obtained.

3D T1-weighted structural data were segmented and processed using FreeSurfer version 7.1.1 (19), to generate the cortical surface to perform surface-based analyses. Proton density images (M0) were registered to the T1 images to generate the co-registration matrix to transform all ASL generated maps to the T1 images in the subject’s space. For the surface-based group analysis, images were co-registered to the Desikan-Killiany template. Z-score maps were calculated for each vertex in the template space. For the epilepsy patients, Z-score measures the deviation of CBF from the average CBF of healthy subjects and normalized by the corresponding standard deviation (SD) for each vertex.

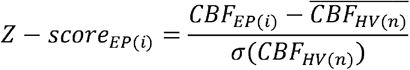

Similarly, the Z-score of healthy subjects was calculated as the deviation of CBF in each subject from the mean of the rest of the healthy controls and then normalized by the corresponding SD.

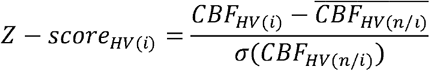

We also calculated the mean of absolute Z-scores (|Z-score|) for healthy and epilepsy subjects.

### Statistical Analysis

To compare age between epilepsy subjects and healthy controls, and epilepsy duration (controlled versus uncontrolled subgroups), the Student T-test was used. For CBF analysis, the mean and SD were calculated for each vertex in the cortical surface in all participants. To compare the CBF derived from single- and multiple-PLD, we calculated Pearson Correlation at each vertex in the cortical surface for each PLD and measured the percentage cortical surface area with significant correlations. The average correlation coefficient was calculated by summarizing all significant correlation coefficients and then normalized by total cortical surface area. The histogram distribution of correlation coefficients at each PLD was also obtained. Significance was defined as P<0.05.

## RESULTS

### Subjects’ Characteristics

Epilepsy group (n=25) included 9 with controlled and 16 uncontrolled epilepsy who have recurrent seizures (>1 year seizure-free) with similar age distribution, age at epilepsy onset, and epilepsy duration (all P>0.05) (Table 1). We did not observe significant differences in perfusion maps between participants with controlled and uncontrolled epilepsy. In addition, our study aimed to explore feasibility and CBF patterns using multiple PLD ASL in focal epilepsy regardless of treatment response. Hence, we combined all epilepsy subjects into one experimental group to increase statistical power. Twenty-two participants had temporal or temporal plus epilepsy, with left foci in 12 (55% of temporal cases), right and bilateral temporal in 8 and 2 respectively (45%). Four participants had extra-temporal foci, with left laterality in 3 out of 4 (Table 1).

**Table 1.** Summary of participants’ characteristics.

|  | Epilepsy | Uncontrolled<br>Epilepsy | Well Controlled<br>Epilepsy | Healthy |
| --- | --- | --- | --- | --- |
| N | 25 | 16 | 9 | 15 |
| (men%, women%) | (48%,52%) | (62.5%,37.5%) | (77.8%,22.2%) | (53.3%,46.7%) |
| Age ± SD (Years) | 41.1±10.5 | 40.8±10.8 | 41.6±10.6 | 35.9±9.5 |
| Age at epilepsy onset | 19.9±12.7 | 20.4±13.0 | 18.9±12.8 | |
| *Seizure focus location* |  |  |  |  |
| Temporal | 21 | 13<sup>&</sup> | 8 |  |
| Extra-temporal | 4* | 3 | 1 |  |
| *2 frontal, 1 parietal, 1broad frontoparietal, &1 temporal plus (with frontopolar involvement) |  |  |  |  |

### Cortical Distribution of CBF in Epilepsy Subjects

A representative CBF_multi_ map and the corresponding T1-weighted structural image are shown in Figure 1A-B. To visualize the global distribution of CBF, we co-registered and mapped the CBF_multi_ values on the brain surface generated from T1-weighted images (Fig 1C). Figure 1 depicts asymmetric CBF_multi_ in a right temporal lobe epilepsy patient (Fig 1A, C). Compared to the left, the right hemisphere showed relatively lower CBF (or left relatively higher CBF).

**Figure 1.**
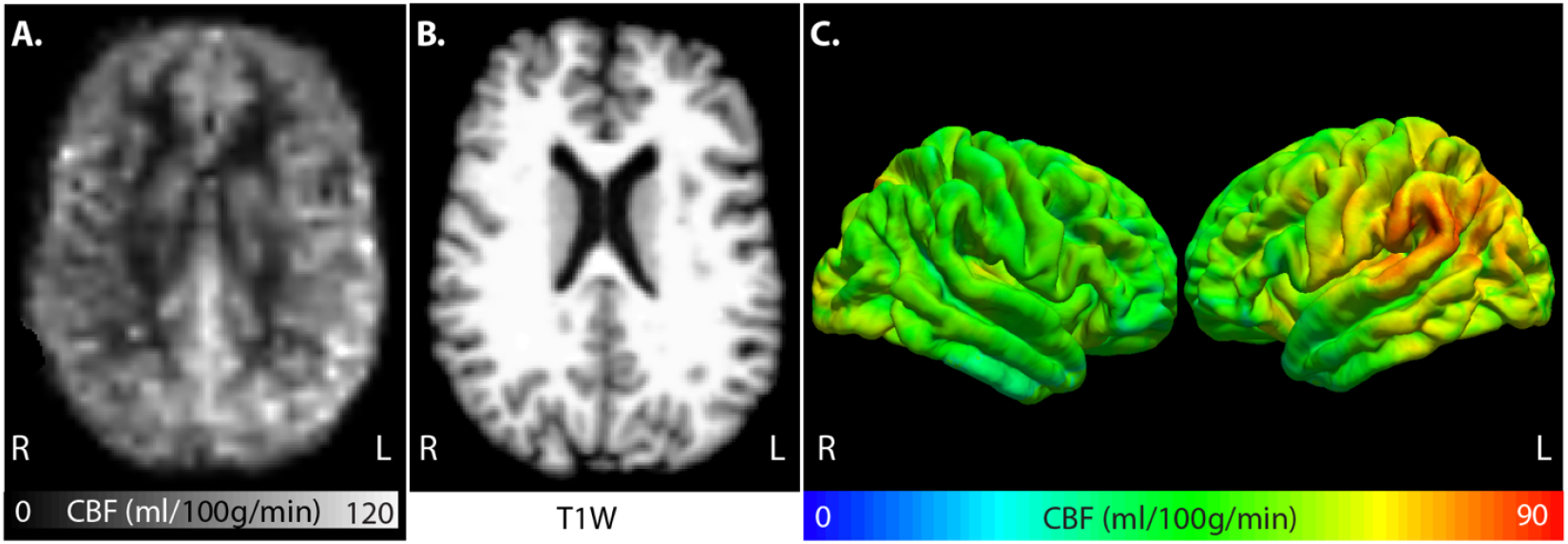
Representative MRI images from a right temporal lobe epilepsy women in her 50’s. The CBF map was derived from multiple post-labeling delay (PLD) images, and it showed higher CBF values on the left side of the brain (A). The T1-weighted structural image indicates the anatomical location of the corresponding CBF map (B). The cortical CBF map was also projected to the pial surface to review the global distribution of CBF values.

To investigate whether the asymmetric CBF is caused by ipsilateral hypo-perfusion or relative contralateral hyper-perfusion, we calculated the CBF Z-scores: CBF_multi_ in patients was normalized to CBF_multi_ in healthy, and CBF_multi_ in healthy was normalized to other healthy subjects.

Figure 2 shows representative cases of Z-score maps with different CBF distribution including focal (Fig 2B), regional (Fig 2A), hemispheric (Fig 2B, E) or global (whole brain, Fig 2C, F) abnormal CBF. Left panel (Cases 1-3) shows examples of hypoperfusion (lower than perfusion in similar regions of healthy participants). Right panel (Cases 4-6) shows examples of hyperperfusion (higher than perfusion in similar regions of healthy participants). Clinical information for each individual is summarized in supplemental table 1.

**Figure 2.**
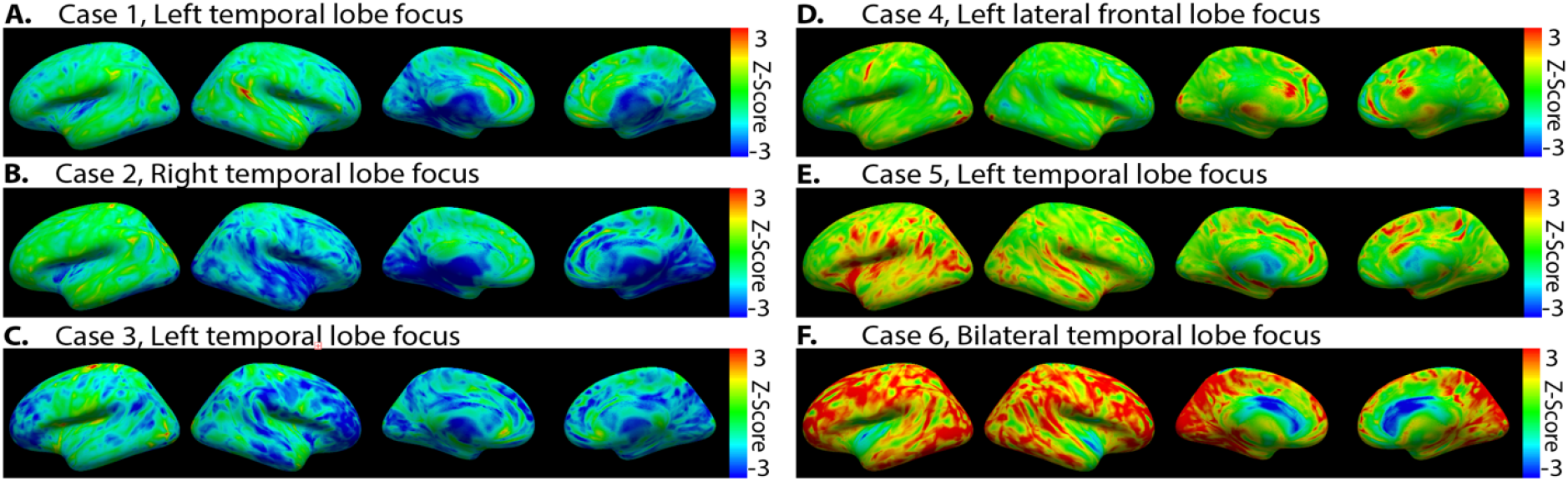
Representative epilepsy cases of Z-score maps. Some subjects showed hypoperfusion (A-C), while other subjects showed hyperperfusion (D-F). Some subjects showed more localized variation of CBF (A, D), some subjects showed lateralized and hemispheric variations of CBF (B, E), and some subjects showed global variations of CBF (C, F).

For group comparison, we measured average absolute Z-scores (|Z-scores|) in healthy (Fig 3A) and epilepsy (Fig 3B) subjects. |Z-scores| measures the degree of deviation from the reference group, whether hypo- or hyper-perfusion, regardless of seizure focus laterality. Compared to the healthy group, the epilepsy group showed larger |Z-scores| across the entire brain cortex (Fig 3). In addition, the left medial temporal lobe in epilepsy group had a larger average |Z-score| compared to the contralateral side or other brain regions.

**Figure 3.**
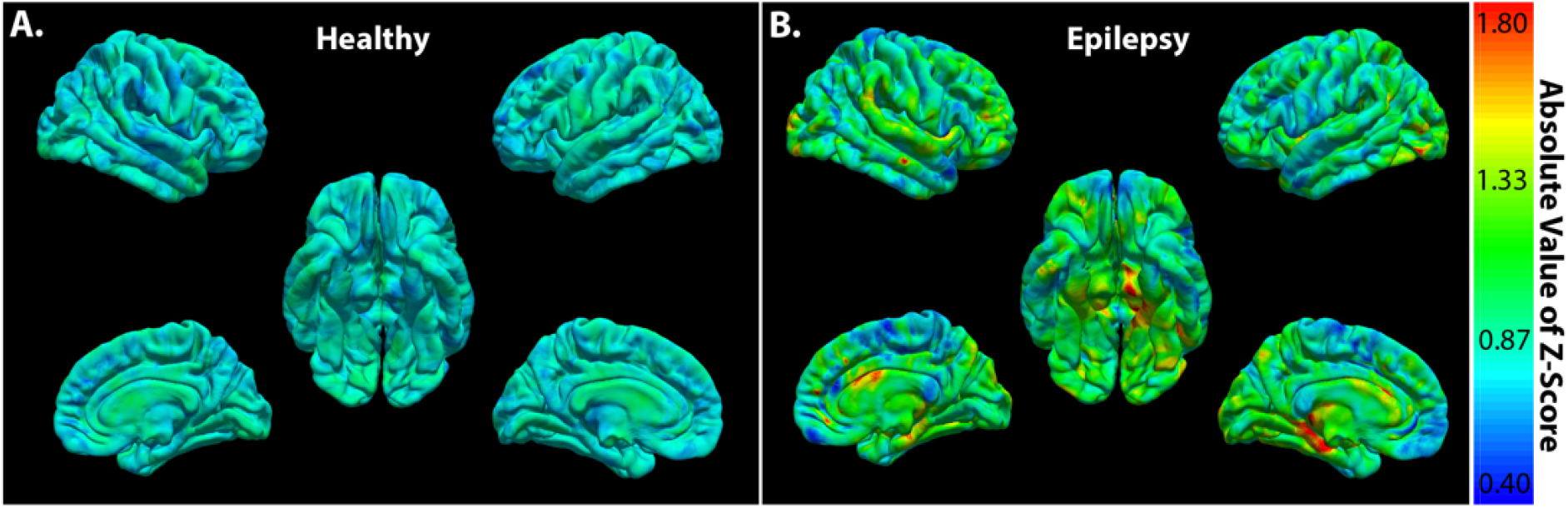
The average map of absolute Z-scores in healthy (A) versus epilepsy subjects (B). Surface voxel-wise analysis maps cerebral blood flow (CBF) onto the cortical surface. This approach aligns perfusion measures with the brain’s folded cortex allowing comparison of localized CBF changes between healthy and epileptic subjects. Compared to healthy controls, the epilepsy group showed larger absolute Z-scores in most brain regions.

### Single vs. Multiple PLD

We used pCASL MRI with multiple PLDs, which provides arrival-time corrected CBF hence generating more reliable measurements, and providing the basis for quantitative analyses without intrasubject (Min-Max) normalization (14). However, adopting multi-PLD pCASL in clinical settings may not be feasible at the current stage because of post-processing and implementation challenges. Therefore, we investigated the correlation of CBF_multi_ and CBF_PLDx_ to evaluate which of the single-PLD parameters provides results as accurate as multiple PLDs measurements. We compared the percentage of cortical area with significant correlation in all participants and within each group. Table 2 shows the percentage of cortical area that showed a significant correlation between the two methods (single/multiple-PLD). Shorter PLD, where signal is mainly arterial, showed the lowest percentage area with good correlation between CBF_multi_ and CBF_PLDx_. As PLD increases, arterial contribution is reduced and increased area showed significant correlations between the two methods (Table 2).

**Table 2.** Comparison between multi-PLD and single PLD ASL MRI shows different percentages of cortical areas with significant correlation, using cortical surface analysis.

| LD (ms) | 650 | 650 | 950 | 650 | 950 | 950 | 2000 | 2000 | 2000 |
| --- | --- | --- | --- | --- | --- | --- | --- | --- | --- |
| PLD (ms) | 200 | 650 | 850 | 900 | 1300 | 1550 | 1800 | 2250 | 2500 |
| All Subject (%) | 48.05 | 94.94 | 99.78 | 99.51 | 100.00 | 99.96 | 99.80 | 99.82 | 98.32 |
| Healthy (%) | 40.06 | 72.45 | 93.79 | 91.14 | 99.12 | 96.41 | 94.91 | 94.31 | 88.36 |
| Epilepsy (%) | 25.43 | 85.83 | 97.70 | 94.43 | 99.52 | 99.14 | 99.17 | 98.93 | 96.21 |

We also summarized all the significant correlation coefficients and normalized by cortical surface area in each group (Table 3). Both comparison methods showed that CBF_PLD1300_ had the optimal correlation (largest areas with significant correlation and highest average correlation coefficients) across our study groups (Table 2,3).

**Table 3.** Average of significant correlation coefficient (normalized by cortical surface area) between single and multiple PLD derived CBF.

| LD (ms) | 650 | 650 | 950 | 650 | 950 | 950 | 2000 | 2000 | 2000 |
| --- | --- | --- | --- | --- | --- | --- | --- | --- | --- |
| PLD (ms) | 200 | 650 | 850 | 900 | 1300 | 1550 | 1800 | 2250 | 2500 |
| All Subject | 0.22 | 0.58 | 0.74 | 0.69 | 0.84 | 0.81 | 0.83 | 0.81 | 0.76 |
| Healthy | 0.26 | 0.51 | 0.72 | 0.68 | 0.87 | 0.84 | 0.83 | 0.81 | 0.74 |
| Epilepsy | 0.13 | 0.55 | 0.72 | 0.65 | 0.82 | 0.79 | 0.81 | 0.80 | 0.74 |

Histogram analysis of correlation coefficient at different PLDs showed that PLDs at 1300, 1550, 1800, 2250 ms generated a more desirable histogram distribution (skew towards 1) as compared to other PLDs (Fig 4).

**Figure 4.**
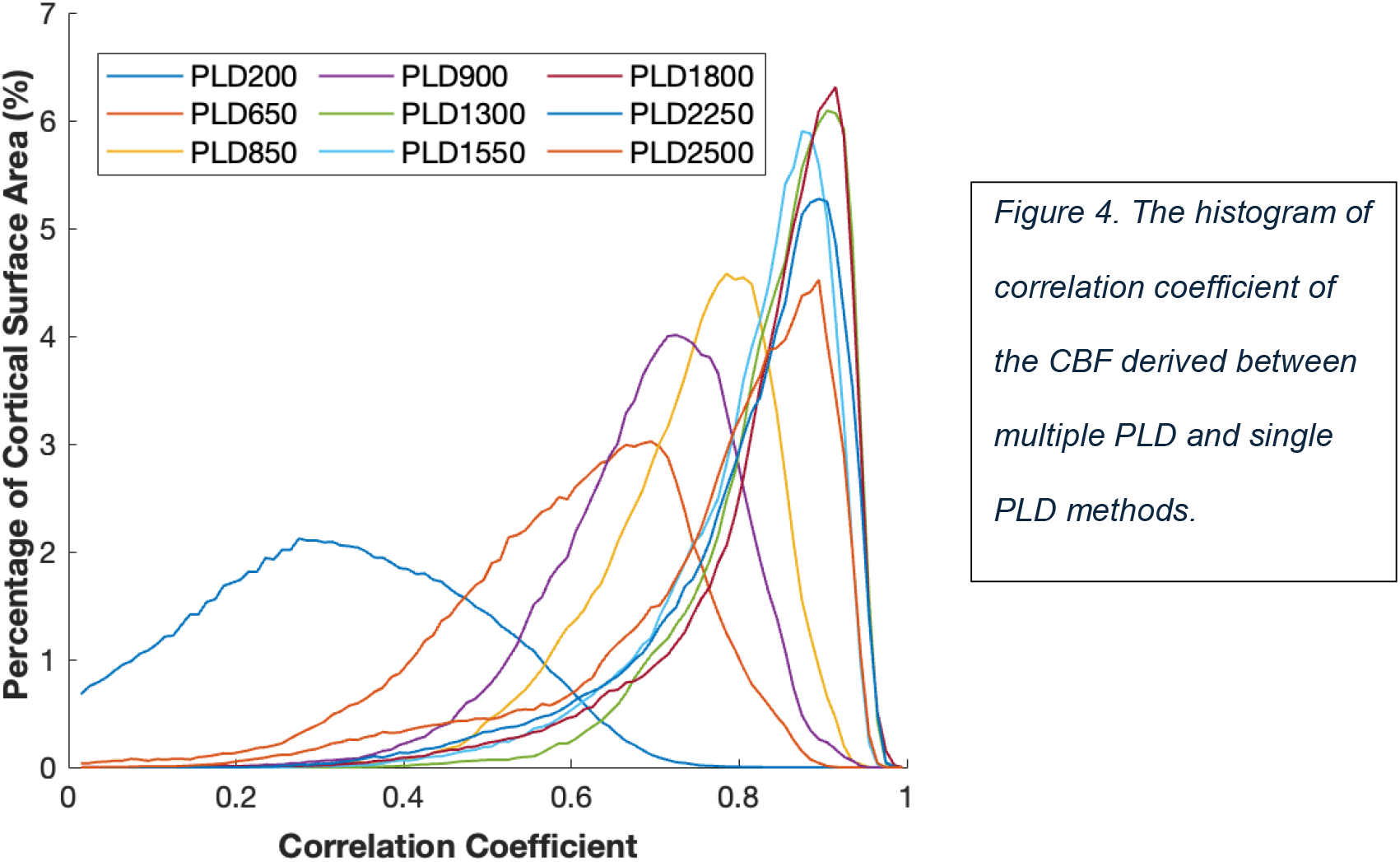
The histogram of correlation coefficient of the CBF derived between multiple PLD and single PLD methods.

To investigate the impact of ATT on CBF quantification using single-PLD, we examined the average ATT and average correlation coefficients at each PLD for all subjects (Fig 5). We did not observe significant differences in ATT between healthy and epilepsy subjects. Hence, we presented the cortical concordance maps combining the two groups. Cortical regions with relatively short ATT (Fig 5B, hollow arrows) showed higher correlation at PLD=1300ms, and the correlation gradually decreases as PLD increases further (Fig 5A, hollow arrows). Meanwhile, in regions with relatively long ATT (Fig 5B, arrow), the correlation coefficient peaked at longer PLDs (Fig 5A, arrows).

**Figure 5.**
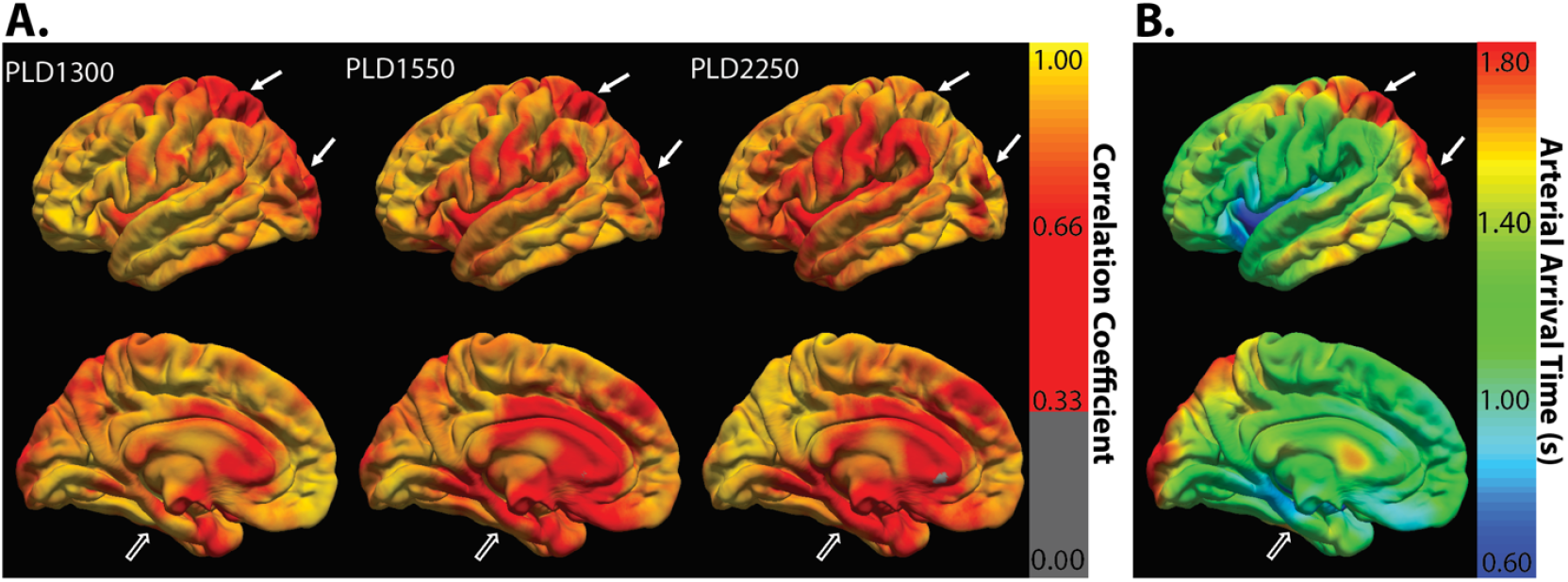
The correlation coefficient maps between PLD and representative single PLD at 1300ms, 1550ms or 2250ms in CBF calculation. Cortical regions with no significant correlation coefficients were labeled gray (A). The correlation coefficients in the inferior brain region (hollow arrows) were higher in PLD at 1300ms and gradually decreased in later PLDs. The same region’s corresponding arterial transit times (ATT) were relatively small (B). Meanwhile, the correlation coefficients in the superior and posterior brain regions (arrows) peaked at a later PLD of 2250ms. These corresponding regions had relatively longer ATT (B).

## DISCUSSION

Seizure focus has been linked to ipsilateral interictal hypoperfusion, yet with variable accuracy (5, 6, 20). In addition, the global patterns of CBF alterations in epilepsy has been less studied. In this work, we observed regional, hemispheric or global variation in interictal CBF in patients with focal epilepsy compared to healthy volunteers. However, this variation may happen in either direction: hypo- or hyper-perfusion. In addition, we observed that the combination of PLD and ATT may affect the reliability of CBF obtained from single PLD pCASL in a spatial-dependent manner.

In epilepsy, whereas ictal hyper-perfusion in seizure focus is well established, interictal CBF generates controversial results (7, 20-26). Interictal hypoperfusion in seizure focus has been observed in previous studies using various techniques, such as SPECT, thermal diffusion flow etry, perfusion MRI, etc. (8-11, 20, 27, 28). However, in 40-50% patients, this hypoperfusion may be discordant with seizure lateralization (20). Several other studies reported interictal hyper-perfusion in seizure focus (7, 29, 30).

Our study investigated the patterns of CBF across the cortex. In line with previous findings, we observed a slightly more dominant perfusion in the left hemisphere of controls (Supplement Fig 1) (8, 11).

This natural lateralization in the healthy condition may contribute to the false localization of seizure focus using asymmetric indices. Therefore, we focused on the deviation of perfusion from controls in each cortical vertex (Z-score). The hyper- or hypo-perfusion was defined by higher or lower CBF values compared to corresponding regions in healthy controls. Ngo et. al. used single-PLD pCASL for CBF surface-based analysis in epilepsy and observed widespread hypoperfusion on average in epilepsy subjects compared to healthy controls (31). The group-wise averaged CBF may mask both temporal and regional variations in CBF between hyper- and hypoperfusion. Therefore, we investigated each subject individually and measured the absolute deviation from healthy controls (absolute Z-scores) to preserve interindividual differences.

We found largest CBF fluctuations in left medial temporal lobe compared to other regions (Fig 3B). It is unclear whether this fluctuation is driven by presence of multiple participants with left temporal foci, soliciting future research in a larger cohort that allows grouping based on seizure focus laterality and focality. In addition to perfusion variability at seizure foci (interictal hyper- and hypo-perfusion), we noted that the pattern of hypo- or hyper-perfusion had spatial variability, ranging from focal (limited to known seizure focus), regional (including broader areas near seizure focus), hemispheric, or even global. Our findings are consistent with the previous postulation that neuronal function and metabolism status might fluctuate in the seizure focus during interictal stage (29, 30). Our findings further suggest that this fluctuation may spread beyond seizure focus. During the “quiet” period between seizures, the so-called interictal period, the brain may still have bursts of epileptiform discharges, corresponding to abnormal neuronal firing, with varying distribution (smaller or broader field) that could affect CBF value (32-35). Therefore, it is plausible that dynamic variability in neuronal activity at seizure focus and connected brain regions modulates CBF and induces CBF swinging during the interictal period, likely reflected in our data due to the long ASL acquisition window (∼21 min) compared to other methods (PET, SPECT).

Blumenfeld et. al. used ictal SPECT and demonstrated broad hyperperfusion during focal impaired consciousness seizure (FIC), whereas involved brain area in focal preserved consciousness seizure (FPC) was more limited (22). Most of our epilepsy participants had FIC (20 out of 25), limiting statistical inferences between the two groups (22). However, we observed the spread of interictal CBF fluctuations in both FIC (n=20) and FPC (n=4) groups. This discordance with the previous finding (22) suggests that abnormally activated and inhibited neuronal network may contribute to widespread CBF variations in epilepsy, yet the involved network or the level of its involvement may be different between ictal and interictal periods.

Multiple-PLD is considered more reliable than single-PLD in CBF measurement (14). However, single-PLD is more feasible in current clinical settings (14). Therefore, we carefully examined the spatial (i.e., brain regions) and temporal (i.e., different PLDs) concordance between single- and multiple-PLD pCASL in CBF calculation. We observed that the spatial concordance varied by a combined influence from the single PLD we chose and the corresponding regional ATT. In the medial temporal lobe, the ATTs were relatively short (Fig 5B), and the correlation peaked at relatively earlier PLD (e.g., 1300ms, Fig 5A). In posterior occipitoparietal area, ATTs were relatively long (Fig 5B), and the correlation peaked at later PLD (e.g. 2250ms, Fig 5A). We presume that this difference in ATT is rooted in different supplying arteries: internal carotid artery territory v.s. posterior cerebral arteries. At longer PLD (i.e., 2500ms), T1 decay and/or labeled blood outflow may be dominant, which may explain its lower correlation.

Our study had several limitations. The labeling efficiency could differ between supplying arteries; contributing to CBF differences between hemispheres. We also adopted the theoretical T1 values of human arterial blood at 3T, not accounting for intersubject T1 variation. Other factors could affect CBF, including caffeine, anti-seizure medications (barbiturates and benzodiazepines), and life habits. We used Hadamard encoding to create 9 perfusion schemes improving SNR. Therefore, our study cannot be matched one-to-one with clinical reports that typically used LD/PLD of 1800 ms for young adults (14). However, the spatial and temporal trends of the correlations between CBF_multi_ and CBF_PLDx_ should be preserved. We also cannot rule out the impact of subclinical seizures on CBF fluctuation. Finally, our pCASL protocol took a total of 21 minutes, that included a T2-preparation module (for BBB exchange). Hence, it would capture the snapshot of average CBF during the 21-minute window and cannot recapitulate transient CBF variations.

## CONCLUSIONS

This study underscores the existence of widespread fluctuations of CBF during the presumably “quiet” interictal period. Our study also suggests caution when interpreting hypo- or hyper-perfusion using normalized, asymmetric or averaged CBF. Finally, when interpreting CBF derived from single-PLD pCASL, caution should be taken given the potential for spatial and temporal variability, and the resulting decreased reliability, particularly in certain brain regions.

## Data Availability

All data produced in the present study are available upon reasonable request to the authors.

## ABBREVIATIONS

CBF: cerebral blood flow
ATT: arterial transit time
pCASL: pseudo-continuous arterial spin labeling
LD: labeling duration
PLD: post-labeling delay
FIC: focal impaired consciousness seizure
FPC: preserved consciousness seizure

## ACKNOWLEDGEMENTS

We are grateful to Dr. Ronald Killiany, our dearly departed colleague, who supported the conception of this study. We thank our participants and the Neurology Clinic who made this work possible.

## DATA SHARING

Data generated or analyzed during the study are available upon request from the corresponding author.

---

**Supplemental Table 1.**
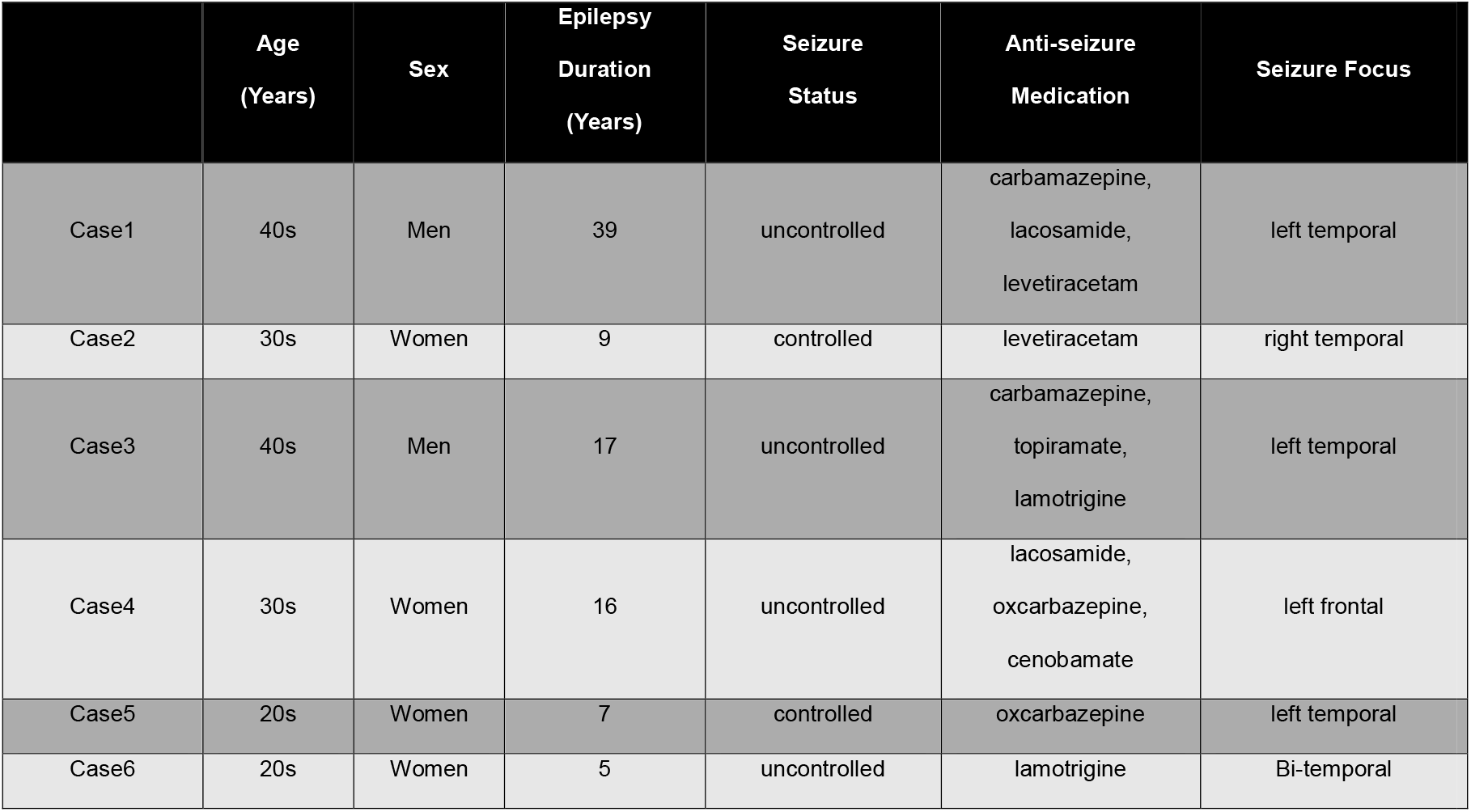
Clinical characteristics of 6 representative cases (in Fig 2) in whom interictal CBF abnormality showed either hypo- or hyper-perfusion on multiPLD ASL.

|  | Age<br>(Years) | Sex | Epilepsy<br>Duration<br>(Years) | Seizure<br>Status | Anti-seizure<br>Medication | Seizure Focus |
| --- | --- | --- | --- | --- | --- | --- |
| Case1 | 40s | Men | 39 | uncontrolled | carbamazepine,<br>lacosamide,<br>levetiracetam | left temporal |
| Case2 | 30s | Women | 9 | controlled | levetiracetam | right temporal |
| Case3 | 40s | Men | 17 | uncontrolled | carbamazepine,<br>topiramate,<br>lamotrigine | left temporal |
| Case4 | 30s | Women | 16 | uncontrolled | lacosamide,<br>oxcarbazepine,<br>cenobamate | left frontal |
| Case5 | 20s | Women | 7 | controlled | oxcarbazepine | left temporal |
| Case6 | 20s | Women | 5 | uncontrolled | lamotrigine | Bi-temporal |

**Supplemental Fig 1.**
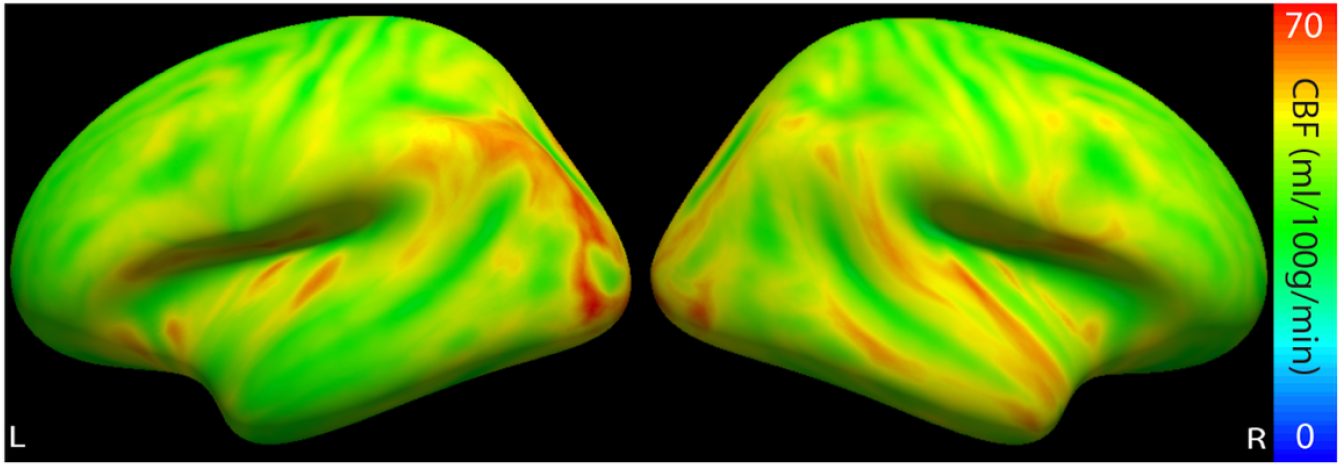
CBF in healthy controls showed left-side dominance.

